# Stage-differentiated modelling of DNA methylation landscapes uncovers salient biomarkers and prognostic signatures in colorectal cancer progression

**DOI:** 10.1101/2020.09.28.20203539

**Authors:** Sangeetha Muthamilselvan, Abirami Raghavendran, Ashok Palaniappan

## Abstract

**Background:** Aberrant methylation of DNA acts epigenetically to skew the gene transcription rate up or down. In this study, we have developed a comprehensive computational framework for the stage-differentiated modelling of DNA methylation landscapes in colorectal cancer. Methods: The methylation β - matrix was derived from the public-domain TCGA data, converted into M-value matrix, annotated with sample stages, and analysed for stage-salient genes using multiple approaches involving stage-differentiated linear modelling of methylation patterns and/or expression patterns. Differentially methylated genes (DMGs) were identified using a contrast against control samples (adjusted p-value <0.001 and |log fold-change of M-value| >2). These results were filtered using a series of all possible pairwise stage contrasts (p-value <0.05) to obtain stage-salient DMGs. These were then subjected to a consensus analysis, followed by Kaplan–Meier survival analysis to explore the relationship between methylation and prognosis for the consensus stage-salient biomarkers.

**Results:** We found significant genome-wide changes in methylation patterns in cancer samples relative to controls agnostic of stage. Our stage-differentiated analysis yielded the following stage-salient genes: one stage-I gene (FBN1), one stage-II gene (FOXG1), one stage-III gene (HCN1) and four stage-IV genes (NELL1, ZNF135, FAM123A, LAMA1). All the biomarkers were hypermethylated, indicating down-regulation and signifying a CpG island Methylator Phenotype (CIMP) manifestation. A prognostic signature consisting of FBN1 and FOXG1was significantly associated with patient survival (p-value < 0.01) and could be used as a biomarker panel for early-stage CRC prognosis.

**Conclusion:** Our workflow for stage-differentiated consensus analysis has yielded stage-salient diagnostic biomarkers as well as an early-stage prognostic biomarker panel. In addition, our studies have affirmed a novel CIMP-like signature in colorectal cancer, urging clinical validation.

## INTRODUCTION

Colorectal adeno-carcinoma (CRC) is a clinically important malignant disease with devastating incidence and mortality, claiming the third spot among all cancers globally, only after lung and breast cancers, and accounting for 1.36 million new cases annually [1]. The etiology of CRC involves chromosomal instability (involving accumulation of mutations in oncogenes and tumor suppressor genes), microsatellite instability (MSI) (leading to loss of DNA mismatch repair) and CpG island methylator phenotype (CIMP), observed in nearly 85%, 15% and 10–40% respectively of all reported sporadic cases [2,3,4].Epigenetic dysregulation is a key driver of these processes, and DNA methylation is the most important epigenetic modification [5,6]. DNA hypomethylation could cause gain-of-function of oncogenes [7], and might aid severe tumor progression [8]. More recently, Timp et al. found that large hypomethylation blocks (hundreds of kb) are a universal characteristic of colorectal cancers and other solid tumors [9]. Hypomethylation could also contribute to tumor initiation and progression by a general increase in genomic instability [10]. DNA hypermethylation could cause loss-of-function of tumor suppressor genes, and hypermethylation in the germline could cause heritable loss of gene expression through genomic imprinting. Aberrant hypermethylation of specific CpG islands has been observed to occur in colorectal cancer. The CIMP was first described in a subset of colorectal cancers in 1999 [11] and later refined to the involvement of five genes *CACNA1G, IGF2, NEUROG1, RUNX3*, and *SOCS1* [12]. Methylation changes contributing to phenotypic aberrations need not be localized to promoter regions but could occur in the gene coding regions and intron-exon structures[13-16].The persistence of such modifications throughout the tumor cell lifetime was demonstrated by Lengauer et al. [17], who showed that methylation aberrations and genome instability were correlated, suggesting a key role for such aberrations in tumorigenic chromosomal segregation processes.

The Cancer Genome Atlas (TCGA) is a comprehensive resource of genome-wide mutation, expression and DNA methylation profiles of 46 different types of cancers [18]. Besides the TCGA, the International Human Epigenetic Consortium (IHEC) is specifically devoted to data-driven understanding of the role of epigenomics in normal vs disease states [19]. Methylation patterns constitute an emerging class of promising prognostic factors mainly due to: (i) the persistence of widespread DNA methylation changes; (ii) the occurrence of such changes much ahead of the consequent changes in gene expression; and (iii) the ability to detect these changes in body fluids and blood plasma [20]. Few methylation markers have been previously translated to clinically applicable biomarkers [21], but it is known that tumor behavior corresponds with differential DNA methylation [80]. Early detection may reduce the mortality rate via tailored adjustments to the treatment regimen, with the result of fewer side-effects and better patient compliance. Chen et al., signalled an era of methylation-based tests by demonstrating an effective screening method to identify multiple types of cancer based on a blood test four years before conventional diagnosis [22]. A consensus approach to identifying significant methylation signatures in each stage of colorectal cancer progression would increase the utility and reliability of putative biomarkers. This motivated our interest in investigating stage-salient DMGs using several model-driven approaches, and evaluating their prognostic significance.

## METHODS

### Data Preprocessing

Processed 27k methylation data (gdac.broadinstitute.org_COADREAD.Merge_methylation _humanmethylation27_jhu_usc_edu_Level_3_within_bioassay_data_set_function_data.Leve l_3.2016012800.0.0.tar) was retrieved from The Cancer Genome Atlas (TCGA) through firebrowse portal (www.firebrowse.org) [23]. The latest clinical data (clinical.cases_selected.tar.gz) was obtained from the GDC data portal (https://portal.gdc.cancer.gov/repository) by matching on patient barcode.

The data containing the methylation β-values for each probe in each sample was converted into a matrix with probes as rows and samples as columns. Each probe corresponds to one CpG site in the genome. A single gene may be under the control of multiple epigenetic sites, hence multiple probes may be associated with the same gene. It is noted that multiple probes usually exist for the same gene. The probes which have “na” values were discarded from the analysis. To transform the range of methylation values from (0,1) to (-∞,+∞), we used the following function on the β-matrix values, to obtain the M-value matrix [24]:

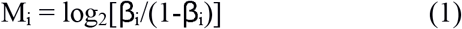

In our study, two M-value matrices were considered: one, where all the probes were used in the analysis; and two, where the probes corresponding to one gene were represented by an average of their values (“averep”), thus reducing the M-value matrix from a probe:sample matrix to a gene:sample matrix. Further, we filtered out the probes/genes showing little change in methylation (defined as σ < 1) across all samples in the M-value matrices. The stages were annotated for both the β-matrix and M-value matrices using the clinical data encoded in the “Pathologic_stage” attribute. Samples with unknown stage (“na” values) were discarded from the analysis. The sample counts in various stages are represented in Table 1.

**Table 1.**
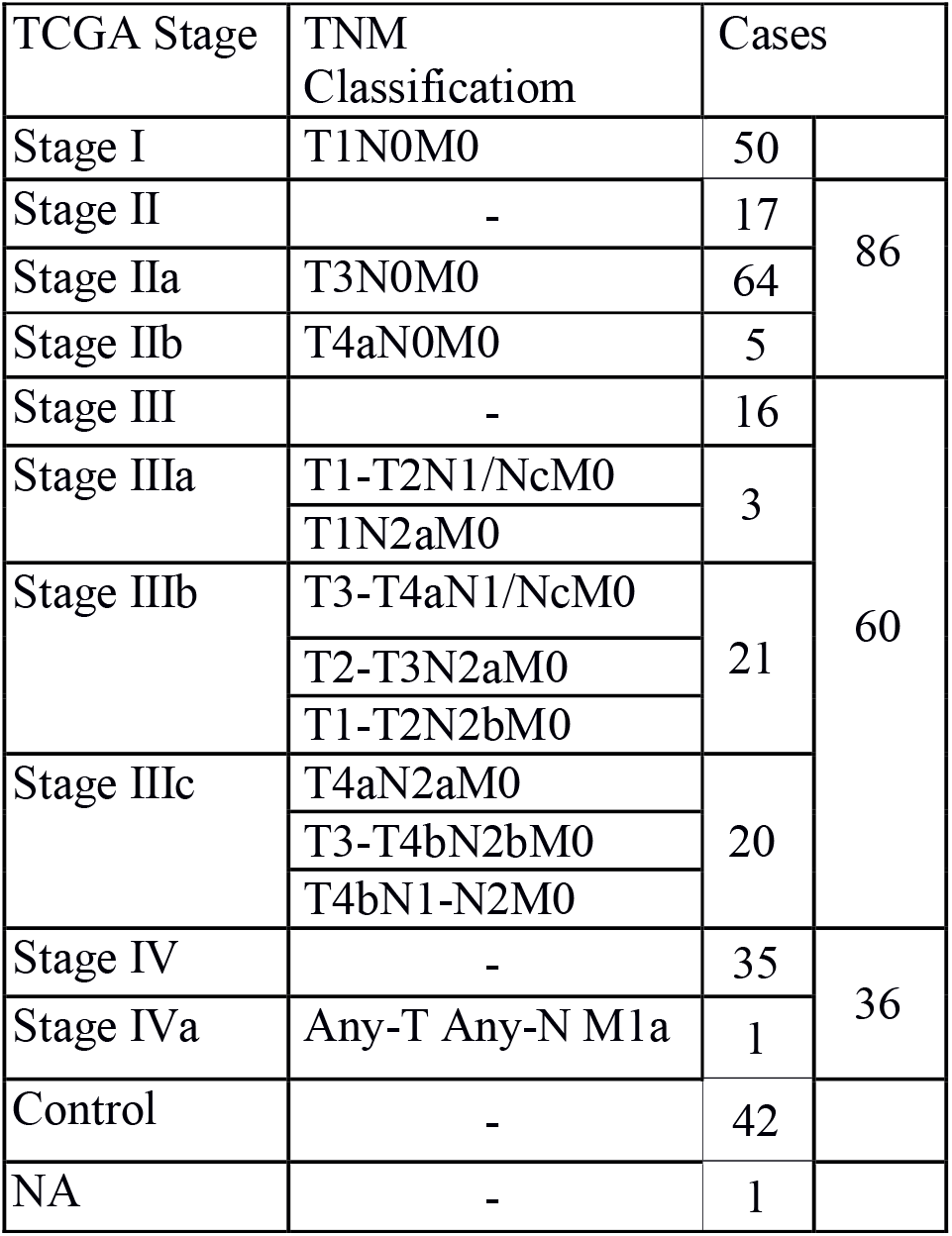
Sample counts in different stages based on 27k methylation COADREAD TCGA data. ‘na’ samples were dropped from analysis, and the sub-stages were combined into the parent stage.

The final β and M-value matrices were subjected to stage-differentiated contrast analysis with a battery of six different methods, described below. All analysis was carried out on R [25].

## Models

### (1) Linear model analysis

Linear modelling is essential to identify linear trends in expression across cancer stages and thereby detect stage-sensitive patterns. We used the R package `limma` [26] for linear modelling of stagewise expression using the complete M-value matrix, with multiple probes per gene (File S1).

### (2) Linear modelling with the averep matrix

This is essentially similar to the above model, except that the input is the averep matrix, where each gene is represented by the average M-value across all its probes (File S2). These alternative representations of the methylation data negotiate a tradeoff with respect to information loss and interpretability. In both the linear models, the control samples contributed to the intercept of the design matrix, while the stages were represented as indicator variables [27]. The linear fit was subjected to empirical Bayes adjustment to obtain moderated t-statistics. These results were then used for the stage-differentiated contrast analysis

### (3) Association between methylation status and phenotype

The strength of the association between the methylation levels of CpG sites and the phenotype of interest (CRC-stage) could enable the identification of relevant markers. We used the R package `CpGassoc` [28] to estimate this association based on ANOVA with multiple hypothesis correction. The β-matrix was used as input, and five factors (control, stage I, stage II, stage III, stage IV) were specified as the target phenotype.

### (4) The Chip Analysis Methylation Pipeline (ChAMP)

The Chip Analysis Methylation Pipeline (ChAMP) integrative analysis suite uses limma to identify differentially methylated probes (DMPs) from the β-matrix [29]. A mapping of sample IDs with the clinical stage phenotype was provided as an additional input file. In addition, the identification of differentially methylated regions (DMRs), consisting of polygenic genomic blocks, was performed using DMRcate in ChAMP (with preset p-value cutoff <0.05) [30]. GSEA was used to identify the enrichment of DMPs and DMRs in the MSigDB pathways [31], using the Fisher Exact test calculation with adjusted p-value < 0.05.

### (5) Modelling expression from methylation

We used the R package `BioMethyl` to model the aggregate expression level of a gene from its methylation patterns [32]. The gene expression matrix was estimated using the methylation β-matrix and then subjected to linear modelling with limma, followed by stage-differentiated contrast analysis.

### (6) Correlation between gene methylation and expression

We used MethylMix2.0 to estimate the correlation between the methylation and actual expression patterns of each gene [33]. The expression data for the samples of interest were retrieved from TCGA (gdac.broadinstitute.org_COADREAD.Merge_rnaseqv2_illuminaga_rnaseqv2_unc_edu_Lev el_3_RSEM_genes_data.Level_3.2016012800.0.0.tar.gz). MethylMix was executed with the preset correlation cutoff (> |0.3|), and statistical significance was assessed using Wilcoxon Rank Sum test with adj. p-value < 0.05.

### Stage-differentiated contrast analysis

A directed two-tier set of contrasts was performed in limma to drill down to the stage-salient genes:

1. Tier I: Stage-differentiated contrast against controls. Four pairwise contrasts were performed, one for each of the stages I, II, III and IV. To identify reliable DMGs, the following criteria were used: |lfc M-value| >2, and adj. p-value <0.001.
2. Tier II: Inter-stage contrasts. Six pairwise contrasts between the stages (namely: I-II, I-III, I-IV, II-III, II-IV, and III-IV) were performed (p-value for each contrast: <0.05).

To illustrate, a putative DMG identified in Tier I would undergo three inter-stage contrasts in Tier II, to ensure stage-salience. For example, a putative stage-II DMG established by Tier I, would have to pass the following inter-stage contrasts: stage-II vs stage-I, stage-II vs stage-III and stage-II vs stage-IV, for confirmation as stage II-salient DMG.

### Identification of stage-salient biomarkers

Finding the consensus of a set of methods with different algorithms overcomes the biases specific to individual methods, and enables screening out false positives. Consensus was obtained by finding the agreement among the results of the various methods used. At least three methods should agree on a given DMG’s stage-salience, for confirmation as *consensus* stage-salient biomarker.

### Survival analysis

The survival data for each patient was obtained from the following attributes encoded in the clinical data: patient.vital_status, patient.days_to_followup, and patient.days_to _death. The association between consensus stage-salient DMGs and case overall survival (OS) was evaluated by univariate Cox proportional hazards regression model using the R `survival` package [34]. This uncovered potential prognostic stage-salient genes from the methylation analysis, using a significance cutoff < 0.05. Such prognostic genes were used as the independent variables in a regression model to estimate the survival risk of each patient. Based on this risk score, patients with colorectal cancer were categorized into high and low groups using the optimal cut point determined by the `maxstat` (maximally selected rank) statistic) [35]. Kaplan-Meier estimation was then applied to the median survival times of these two groups for flagging significant differences, providing prognostic assessment of the biomarkers of interest.

## RESULTS

### Linear modelling at the probe-level

The number of significant genes present in each stage-control pair from the Tier-I contrasts is shown in Figure 1(i). Using the top 100 DM genes of the linear model (given in Supplementary Information S3), we found a clear separation between controls and stage samples (Figure 1(ii)). The top genes in each stage (by adjusted p-value of contrast with control) are shown in Table 2, with |lfc M-value| and inferred regulation status. Figure 2 shows boxplots of stagewise methylation levels for two representative genes: (1) TMEM179, mutations in which could cause MSI [36]; and (2) MEOX2 whose promoter methylation status is a known CRC marker [37].The top four genes of each stage were used to construct a stagewise methylation heatmap(Figure 3). The stagewise methylation patterns of the top five linear model genes are also shown, in Figure 4. It is notable that a naturally occuring read-through fusion protein GPR75-ASB3 is the top linear model gene with significant differential expression in all stages relative to the control. GPR75-ASB3 is positively differentially expressed in the lung as well as different keratinocyte cell types, and evidence is emerging of its role in other cancers [79]. In this light, GPR75-ASB3 could play a significant role in colorectal cancers which are of epithelial origin. The top 100 significant stage-specific genes, listed in S3, were used in the consensus analysis.

**Table 2.** Top ten genes of the linear model at the probe level. The log fold-change of M-value of the probe in each stage relative to the controls, followed by p-value adjusted for the false discovery rate, and the methylation status of the gene in the cancer stages with respect to the control

| ID | StageI | StageII | StageIII | StageIV | adj. p-val | Methylation status |
| --- | --- | --- | --- | --- | --- | --- |
| GPR75-ASB3 | 2.280584 | 2.190314 | 2.159852 | 2.320709 | 2.90E-82 | Hyper |
| TM4SF19 | -3.62705 | -3.57631 | -3.72308 | -3.71059 | 4.08E-82 | Hypo |
| CNRIP1 | 2.743322 | 2.605539 | 2.678591 | 2.974642 | 5.57E-78 | Hyper |
| PDE4A | 1.682123 | 1.578549 | 1.599996 | 1.707521 | 1.27E-71 | Hyper |
| KRTAP11-1 | -2.36005 | -2.29961 | -2.37583 | -2.39962 | 9.04E-70 | Hypo |
| ADHFE1 | 3.153667 | 2.967236 | 3.000252 | 3.432456 | 2.03E-69 | Hyper |
| FAM123A | 3.56288 | 3.181145 | 3.429552 | 3.895594 | 5.54E-69 | Hyper |
| KHDRBS2 | 2.302345 | 2.16154 | 2.103098 | 2.339009 | 4.41E-68 | Hyper |
| AJAP1 | 2.528654 | 2.438316 | 2.462037 | 2.6367 | 9.36E-68 | Hyper |
| NALCN | 2.959115 | 2.796519 | 2.949049 | 3.250277 | 9.36E-68 | Hyper |

**Figure 1.**
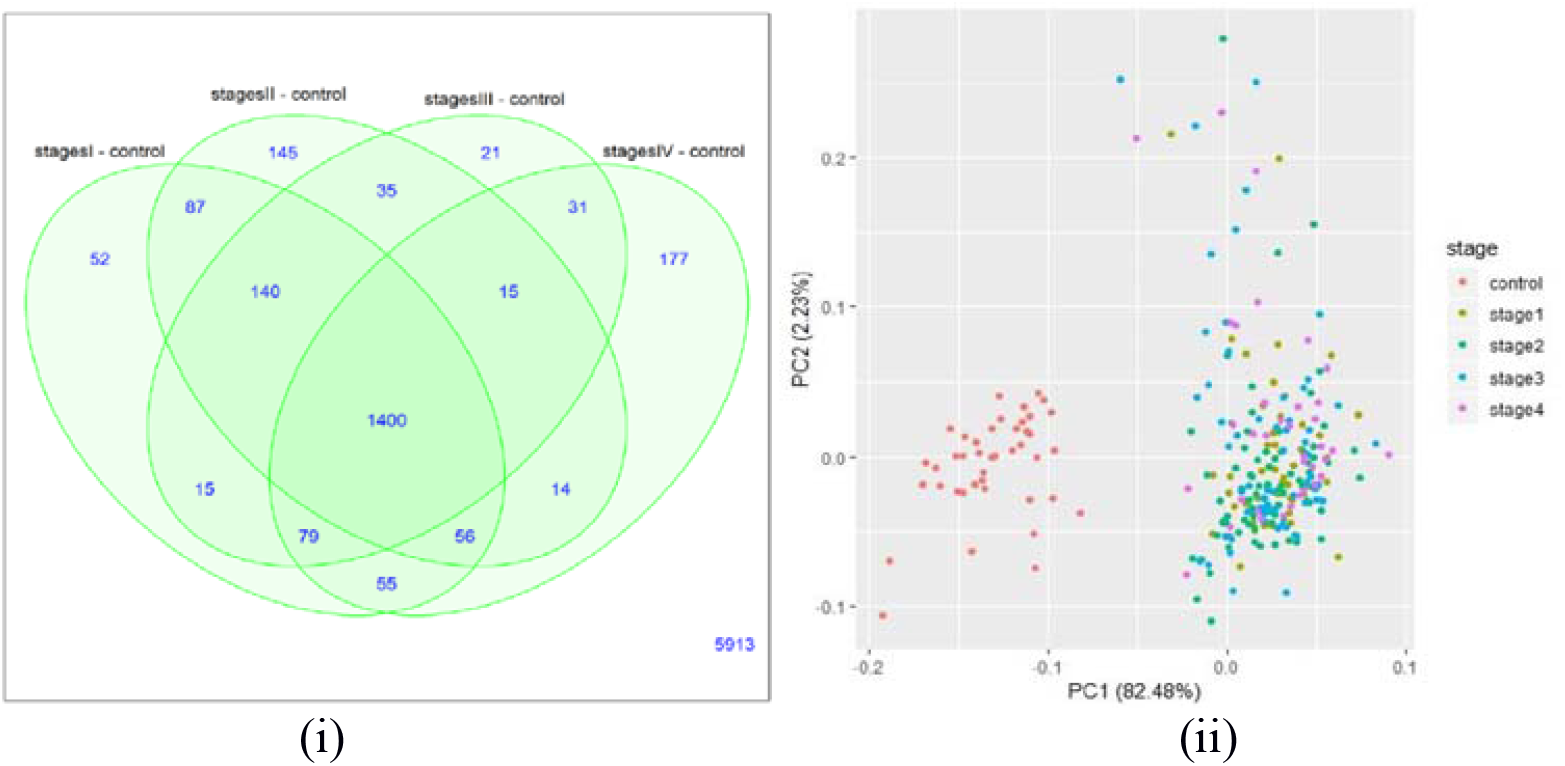
Linear modelling with M-value matrix, all probes. (i) Distribution of number of significant genes in each stage relative to the control. (ii) Sample distribution obtained by plotting the first two principal components for the top 100 genes. A clear separation of controls and cancer samples (labelled by stage) could be seen.

**Figure 2.**
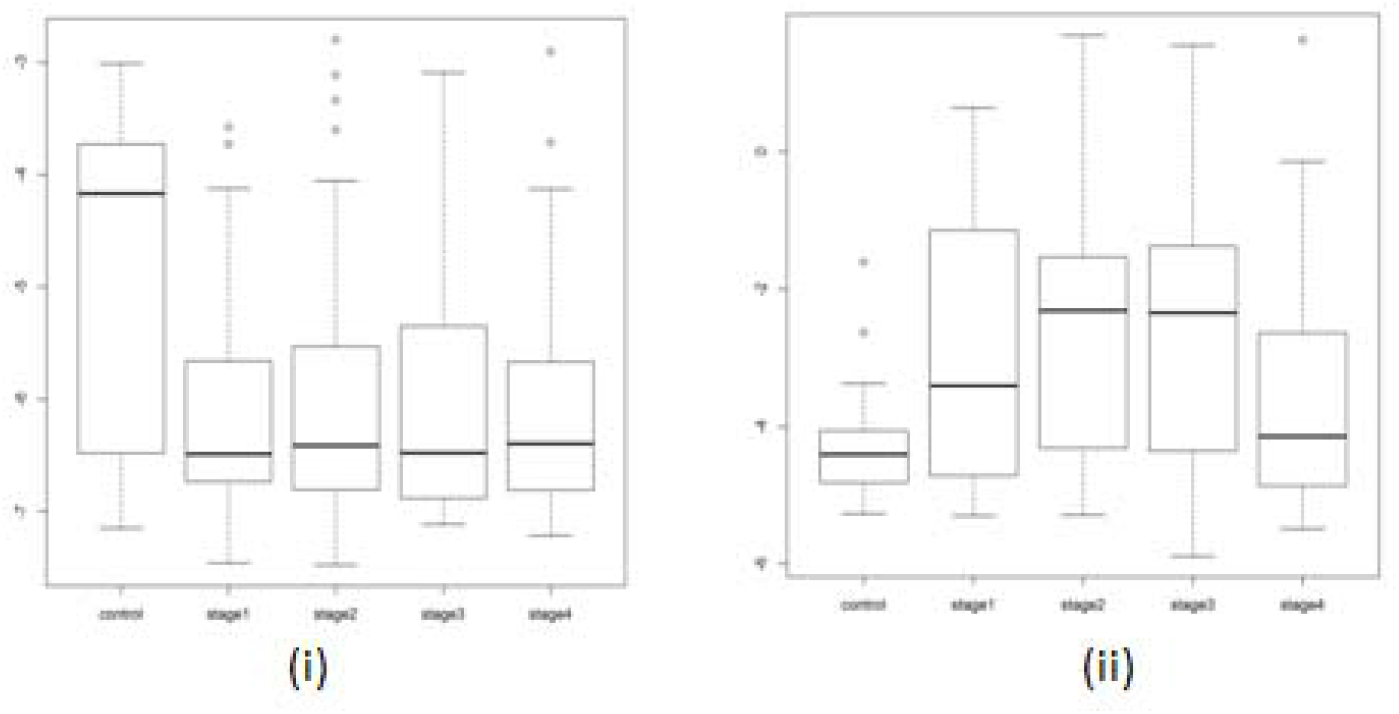
Stagewise methylation levels of differentially methylated genes. (i) TMEM179 (ii) MEOX2

**Figure 3.**
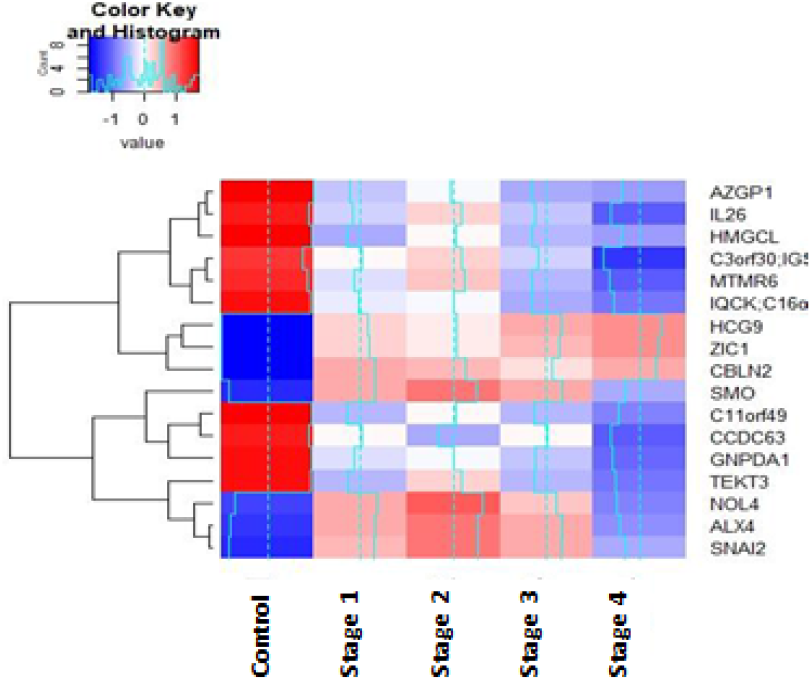
Stagewise methylation portrait using the top 4 significant stage-specific DMGs identified from linear model at the probe level. The contrast with the control is especially evident.

**Figure 4.**
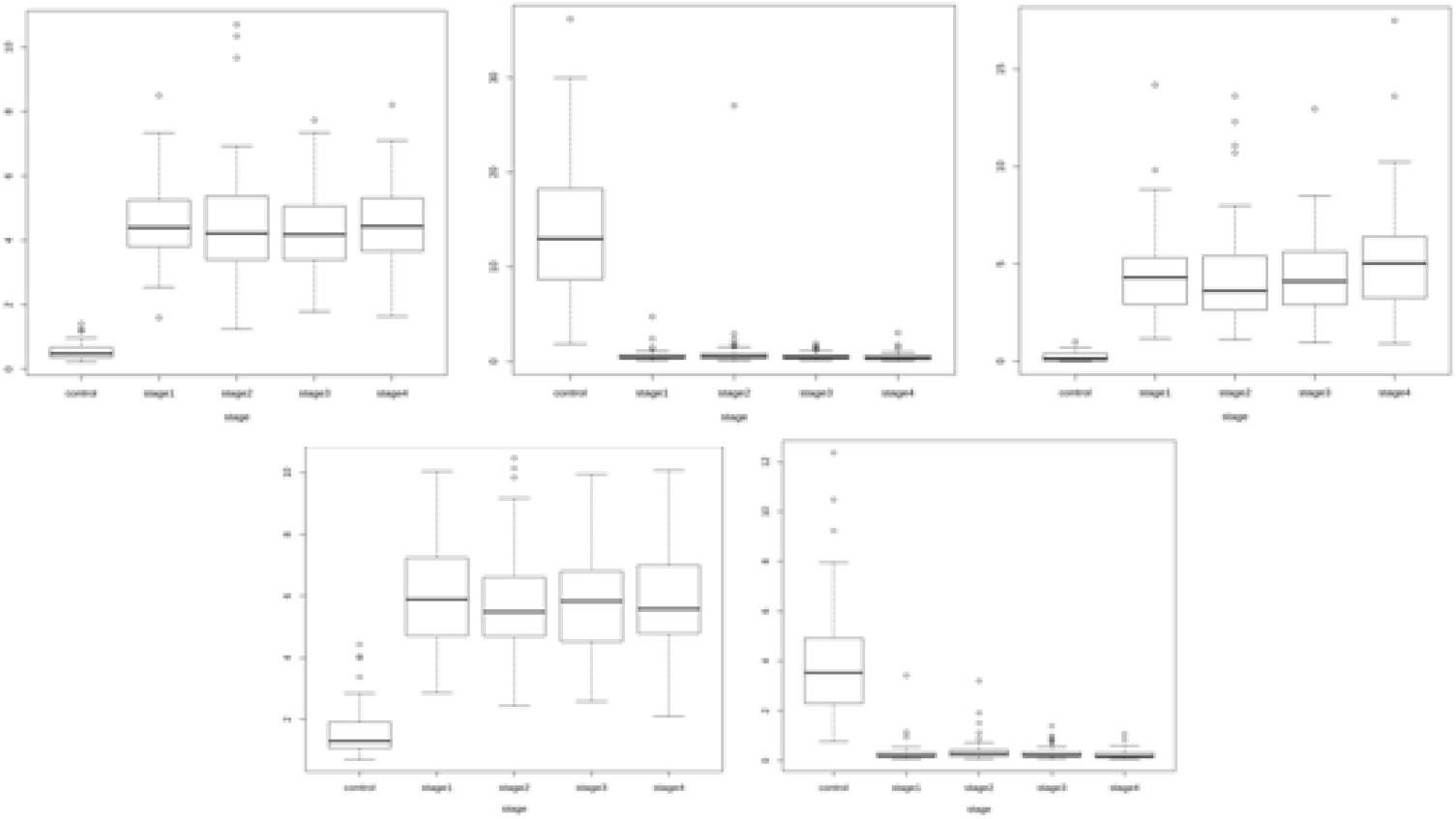
Top 5 DMGs of the full linear model: (i) GPR75-ASB3, (ii) TM4SF19, (iii) CNRIP1, (iv) ADHFE1 and (v) KRTAP11-1. For each gene, notice that the trend in methylation could be either hyper-or hypo-methylation relative to the control. In particular, TM4SF19 and KRTAP11-1 are hypomethylated whereas CNRIP1, GPR75-ASB3, PDE4A are hypermethylated.

### Linear modelling at the gene-level (averep)

The genes with more than one probe were averaged to a single methylation value, which was then further analyzed. The number of genes present in each stage-control pair from the Tier-I contrasts is shown in Figure 5(i). Using the top 100 genes of the linear model (given in Supplementary Information S4), we found a clear separation between controls and stage samples (Figure 5(ii)). The top genes in each stage (by adjusted p-value of contrast with control) are shown in Table 3, with |lfc M-value| and inferred regulation status. Figure 6 shows the boxplots of stagewise methylation levels for two representative genes, NALCN and GLRX. Mutations in NALCN have been reported in sporadic CRC [38]; here NALCN is seen to be significantly hypermethylated, indicating the same outcome (loss of function) could be effected in multiple ways. GLRX is a target of the activating transcription factor MEOX2 [39]. The top four genes of each stage were used to construct a stagewise methylation heatmap (Figure 7). The stagewise methylation patterns of the top five linear model genes are also shown, in Figure 8. It is observed that *LY6H* showed both hypermethylation and hypomethylation when compared to the control samples, indicating the role of experimentation necessary to clarify its role in colorectal cancer progression. The top significant 100 genes of each stage, listed in S4, were used for the consensus analysis.

**Table 3.** Top ten genes of the linear model at the gene level, using average values of methylation. The log fold-change of M-value of the gene in each stage (relative to the control) is given, followed by p-value adjusted for the false discovery rate and the methylation status of the gene in the cancer stages with respect to the control. A consistent methylation pattern is observed for all the top genes.

| ID | StageI | StageII | StageIII | StageIV | adj.P.Val | Methylation status |
| --- | --- | --- | --- | --- | --- | --- |
| TM4SF19 | -3.62512 | -3.576 | -3.72406 | -3.70707 | 1.01E-82 | Hypo |
| GPR75-ASB3 | 2.279406 | 2.186388 | 2.154401 | 2.319774 | 7.39E-82 | Hyper |
| CNRIP1 | 2.742948 | 2.602632 | 2.673634 | 2.97448 | 2.74E-77 | Hyper |
| KRTAP11-1 | -2.35972 | -2.30206 | -2.37938 | -2.3988 | 3.65E-70 | Hypo |
| ADHFE1 | 3.152625 | 2.962694 | 2.994224 | 3.4316 | 4.02E-69 | Hyper |
| FAM123A | 3.561976 | 3.177355 | 3.423787 | 3.894354 | 1.01E-68 | Hyper |
| AJAP1 | 2.527459 | 2.435239 | 2.458005 | 2.635741 | 3.21E-67 | Hyper |
| NALCN | 2.957453 | 2.792623 | 2.943573 | 3.248803 | 1.07E-65 | Hyper |
| IRF4 | 1.98933 | 1.824373 | 1.887686 | 2.128157 | 1.07E-65 | Hyper |
| PRKAR1B | 3.381341 | 3.131659 | 3.244301 | 3.494884 | 1.07E-65 | Hyper |

**Figure 5.**
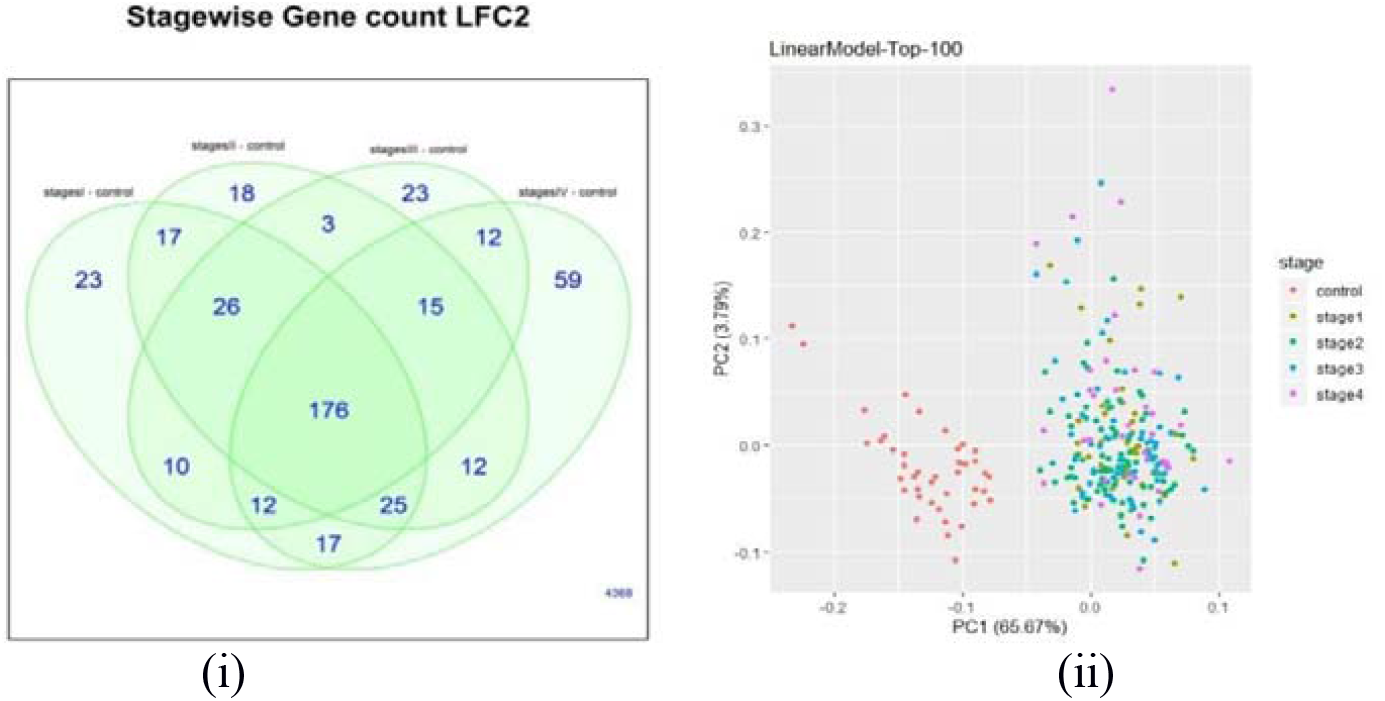
Linear modelling with M-value matrix, averep.(i) Distribution of number of significant genes in each stage relative to the control. (ii) Sample distribution obtained by plotting the first two principal components of the top 100 genes from the linear model. A clear separation of controls and cancer samples (labelled by stage) could be seen.

**Figure 6.**
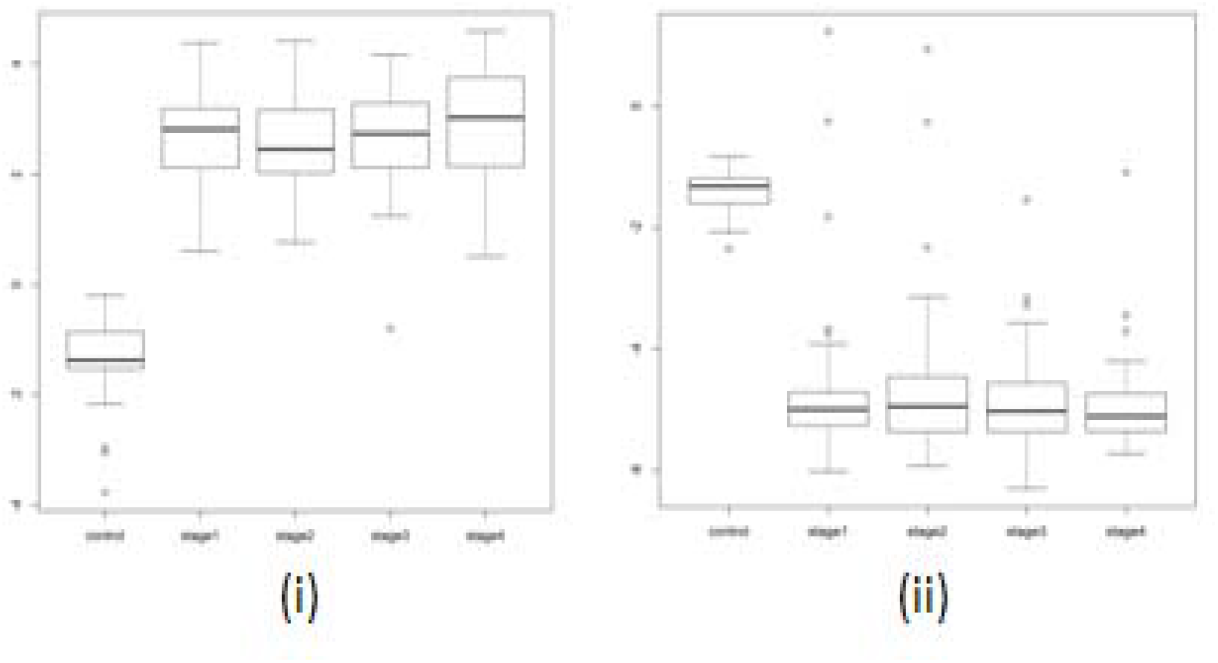
Stagewise methylation levels of differentially methylated genes from averep analysis: (i) NALCN, and (ii) GLRX.

**Figure 7.**
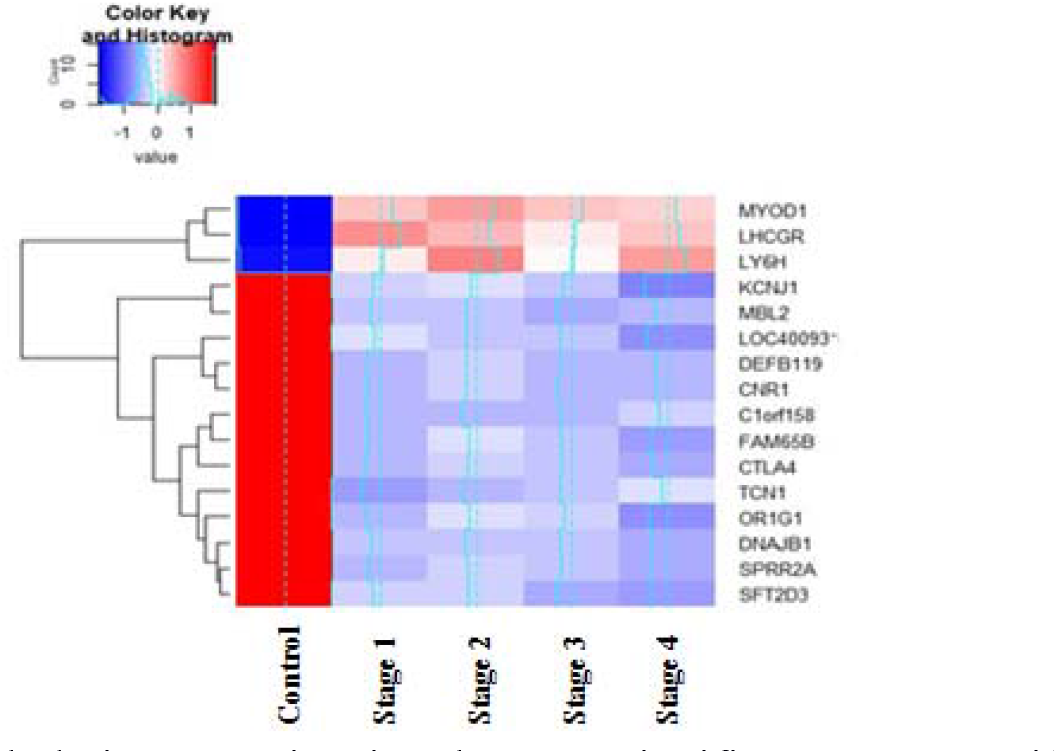
Stagewise methylation portrait using the top 4 significant stage-specific DMGs identified from linear model at the gene level. The contrast with the control is especially outstanding.

**Figure 8.**
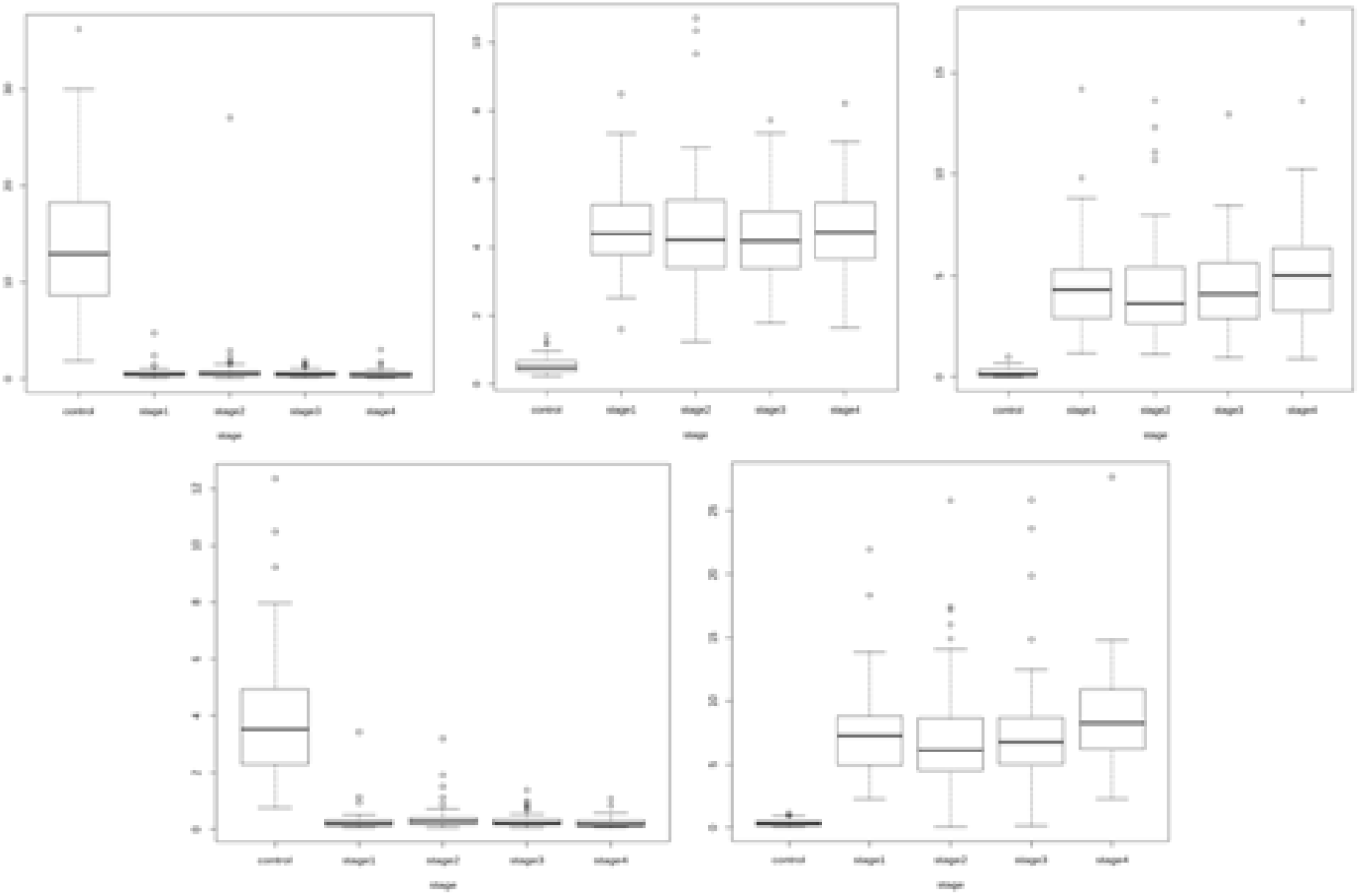
Boxplots of top 5 linear model genes. For each gene, notice that the trend in expression could be either hyper- or hypo-methlation relative to the control. In particular, TM4SF19 and KRTAP11-1 are hypomethylated whereas GPR75-ASB3, CNRIP1, ADHFE1 are hypermethylated.

### Association with phenotype

The ANOVA from CpGassoc yielded p-values and log fold-changes, which were used to identify significant genes for each stage using the criteria given in Methods (Figure 9). The top 100 genes of each stage from this analysis (given in Supplementary Information S5) were used for the consensus investigation.

**Figure 9.**
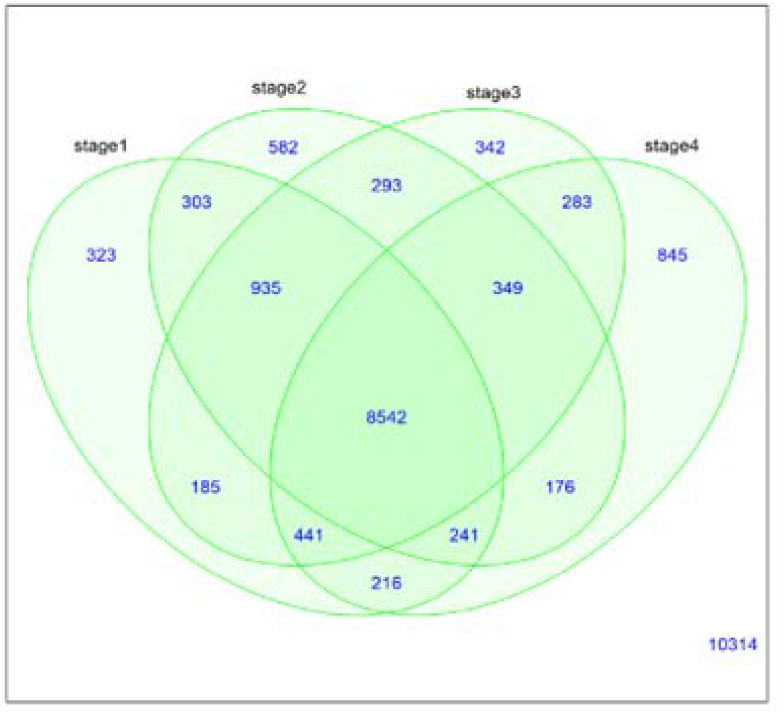
Venn diagram for CpG association analysis showing the distribution of number of significant genes in each stage relative to the control.

### DMP analysis with ChAMP

The summary features of the β matrix dataset were evaluated using ChAMP (Figure 10). The DMPs were identified using CHAMP analysis from the β matrix. All the inter-stage contrasts yielded null results (i.e, no significant genes), except for stageII – stageIV contrast. Due to this, the top 100 DMPs from the stage vs control contrasts were used for the consensus analysis directly. Contrasts that showed significant DMPs were subjected to a further DMR analysis, to enable identification of DM genes. The stage-salient DMR regions (genes) determined are provided in Supplementary Information S6, and summarized in Table 4. The stage-II vs stage-IV DMR contrast yielded three genes, namely PLAG1, SOCS2, and NNAT. It is observed that these genes might be critical players in the transition to malignancy. Interestingly, some genes were differentially methylated in all the stagewise contrasts with the control; such genes are differentially methylated agnostic of stage, and could serve as valuable drug targets for CRC therapy. The top such genes included EYA4, WT1, DCC, RP11, GATA4, MSX1, DLX5, BNC1, WT1-AS, and ZIM2. A total of 31 such genes were identified and tabulated in Supllementary Information S7. The DMPs and DMRs from the analysis were subjected to GSEA and these results could also be found in Supplementary Information S6. Figure 11 shows representative DMP and DMR plots using MethylMix.

**Table 4.**
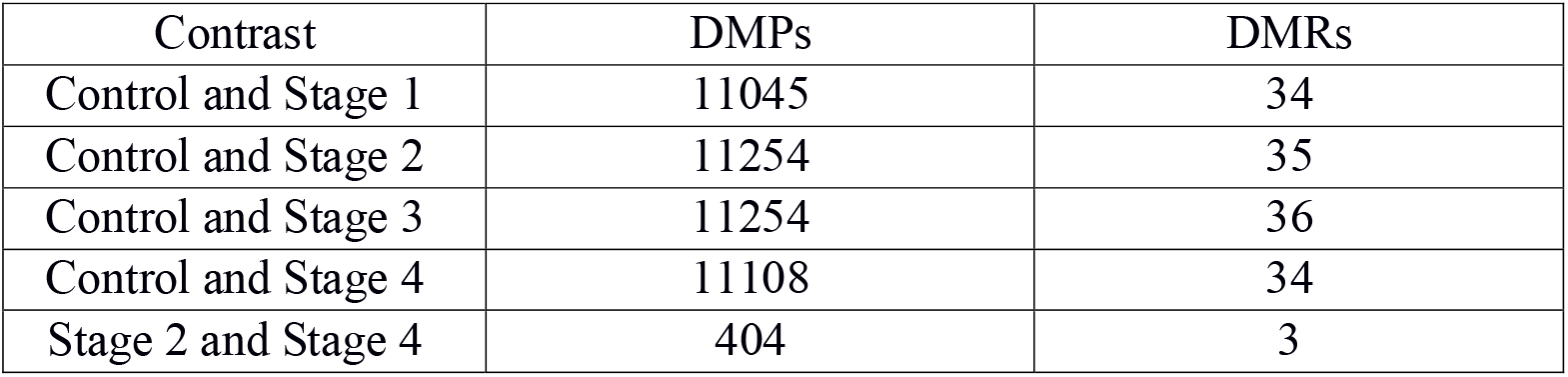
Contrast-wise counts of DM probes and DM regions.

**Figure 10.**
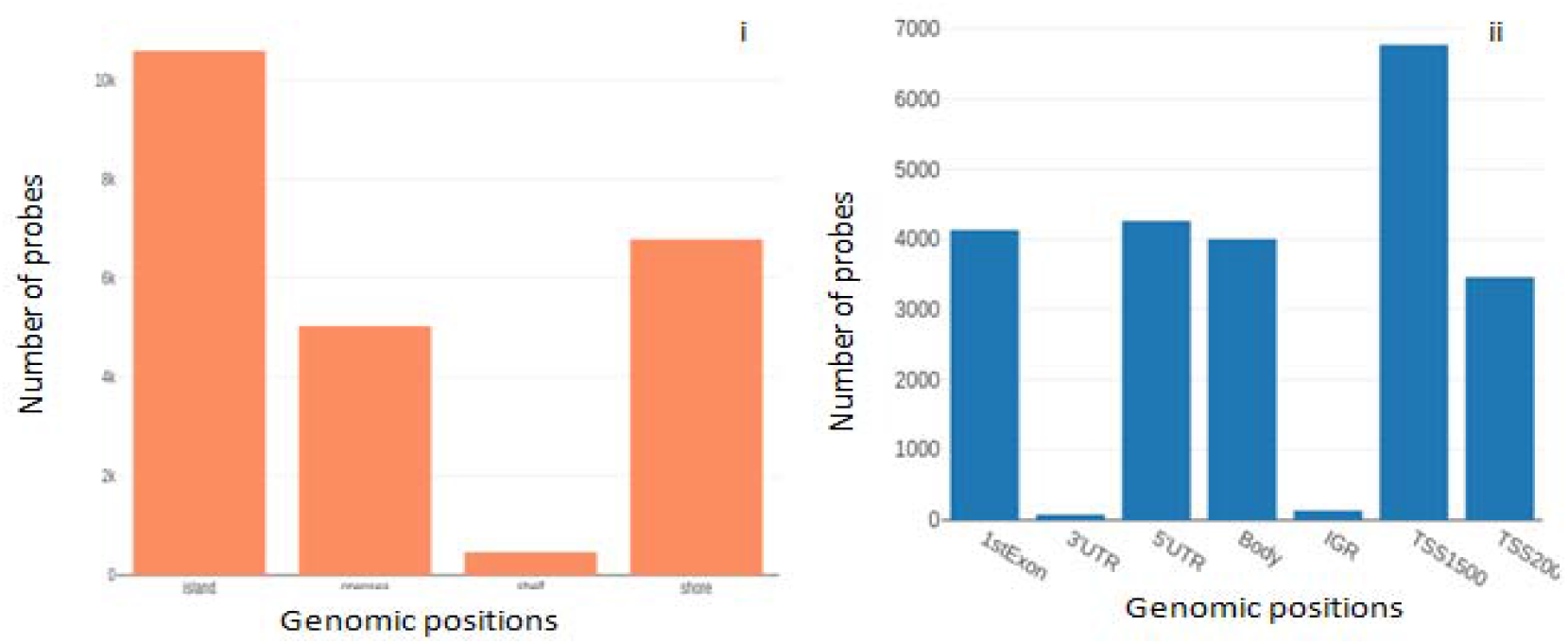
Distribution of probes based on (i) Genomic position: opensea, shore, island, shelf; (ii) gene context: transcription start site (TSS), exons, un-transcribed regions (UTRs), and inter-genic regions (IGR).

**Figure 11.**
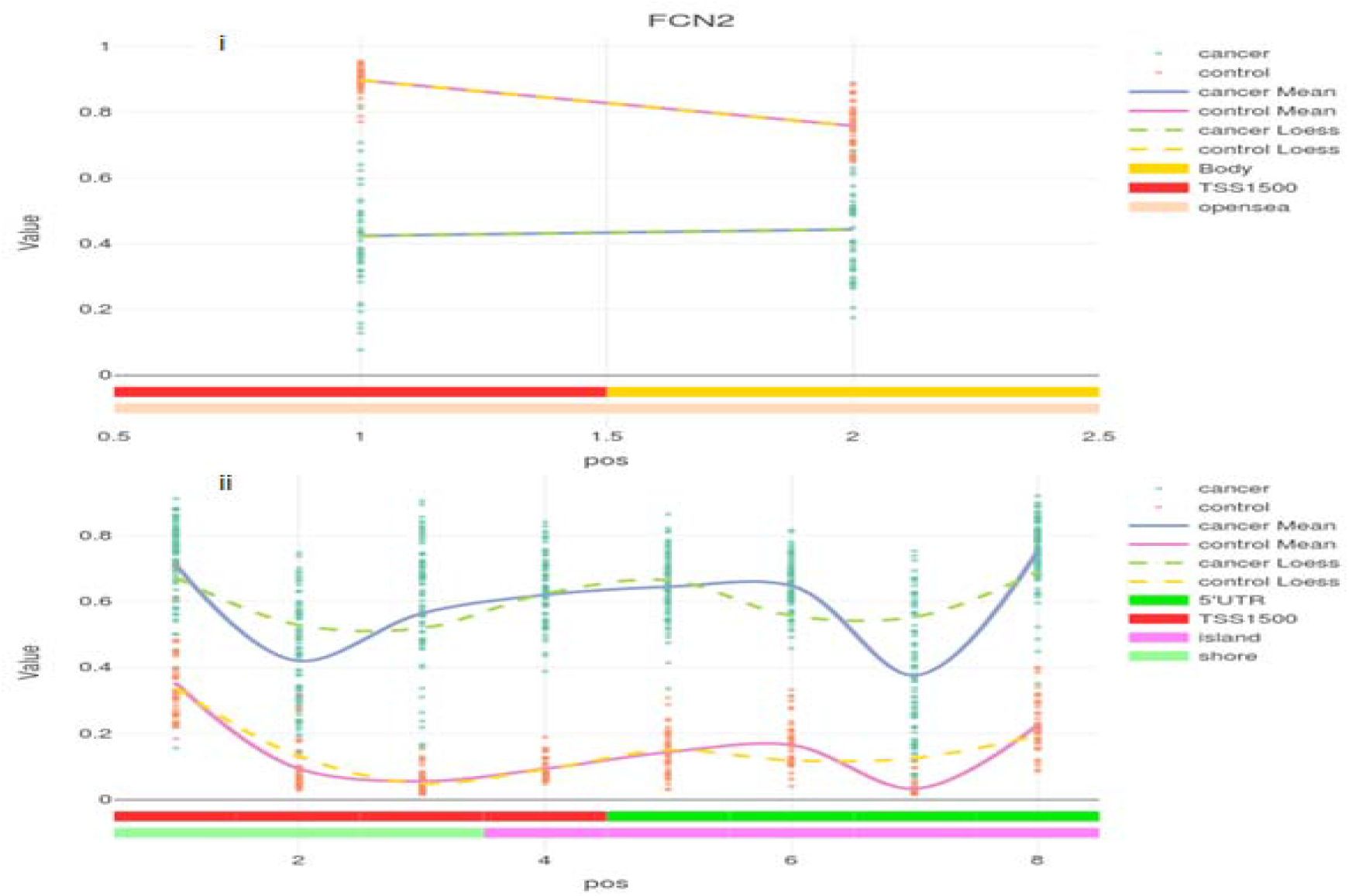
(i) DMP plot of FCN2 for stage-I vs control illustrating significant hypomethylation (ii) DMR plot of transcriptional activator EYA4 for stage-I vs control illustrating significant hypermethylation. Solid lines represent mean values while dashed lines represent the loess.

### Methylation and Gene Expression Correlation analysis

Mixture models of genes, indicative of the number of methylation states, were constructed using MethylMix, and the top three genes from an overall cancer vs control comparison are shown in Figure 12. The estimated correlation between the methylation levels and actual gene expression for the same genes is depicted in Figure 13. Genes were differentially methylated and designated as ‘driver’ genes if the p-value of the contrast being studied was significant. The calculated differential methylation (DM) values from stage vs control contrasts ranged from −0.7 to +0.8, and genes were classified as hyper- or hypo-methylated based on the DM value. There were 209, 441, 275, and 134 driver genes in each of the contrasts with the controls (stage-I, stage-II, stage-III and stage-IV, respectively). All between-stages contrasts yielded null DM genes. The results from this analysis, including driver genes for all the contrasts, are provided in Supplementary Information S8. Top 100 genes from each comparison were taken forward for the consensus analysis. Certain genes emerged common to all the four comparisons, indicating stage-agnostic differential methylation events. The top such genes included *CCDC88B, C1orf59, CHFR, ZP2, HOXA9, ELF5, FAM50B, MUC17, TBX20*, and *VSIG2. S*tage-agnostic genes hold promise as therapeutic targets for the treatment of colorectal cancer; the complete list of 56 stage-agnostic genes arising out of the MethylMix analysis is provided in Supplementary File S9.

**Figure 12.**
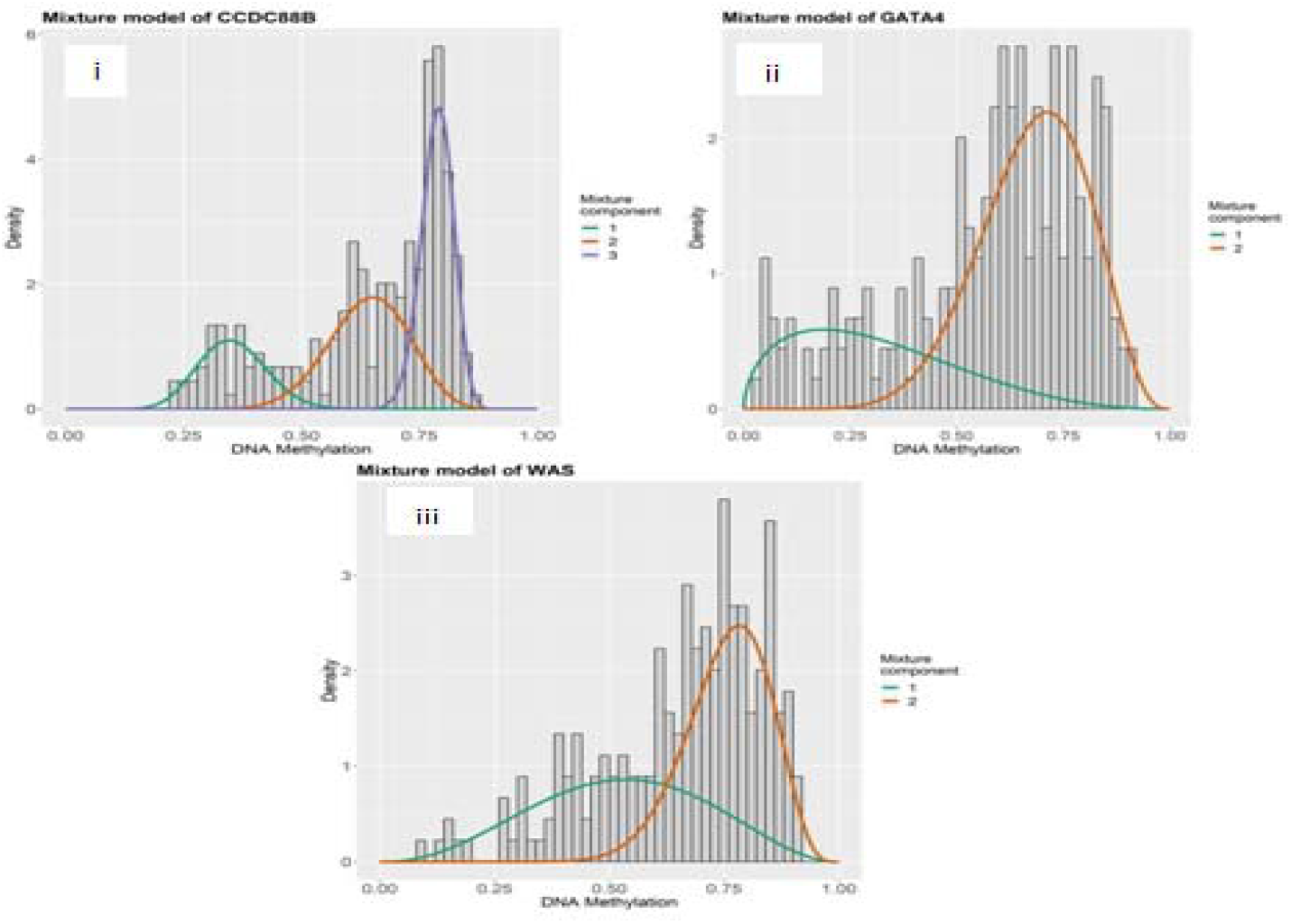
Mixture models of the genes GATA4, CCDC88B, and WAS. The x-axis indicates the degree of methylation; the y-axis represents the frequency of that particular methylation degree; and the mixture component curves represent density fits of the histogram.

**Figure 13.**
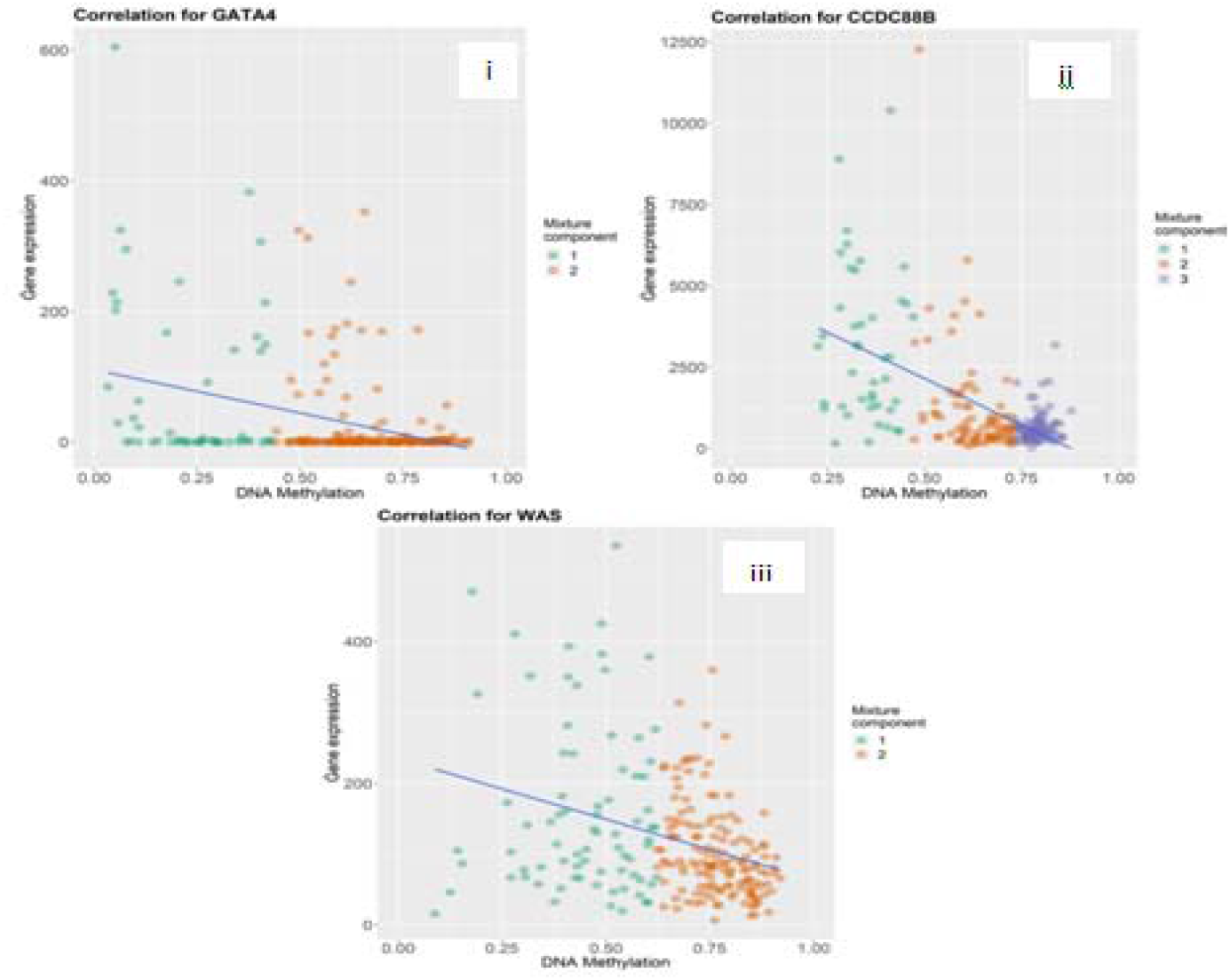
Correlation plots for i. GATA4, ii. CCDC88B, iii. WAS. A negative correlation between methylation and expression is evident, indicating that methylation acts to repress gene transcription, though the strength of the inverse correlation varies from gene to gene. Colour indicates the mixture model fit (cf. Fig. 12).

### BioMethyl analysis

The significant stage-specific DEGs identified by this BioMethyl are shown in Figure 14. Top 100 genes of each stage from this analysis were taken for consensus analysis. The stage-specific genes from this analysis are presented in the Supplementary Information S10.

**Figure 14.**
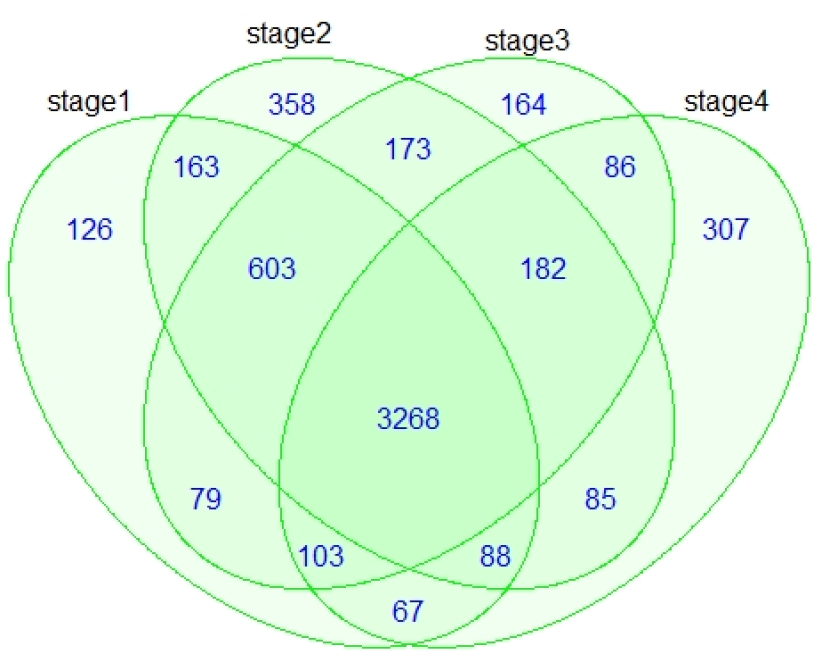
Venn diagram for BioMethyl-based Gene expression modelling showing the distribution of number of significant genes in each stage relative to the control.

### Stage-salient consensus biomarkers

The top 100 significantly differentially-expressed genes of each stage from all the methods discussed above (collated in Supplementary Information S11) were used for the consensus determination. The consensus analysis yielded seven stage-salient DMGs: one stage-I gene *(FBN1)*, one stage-II gene *(FOXG1)*, one stage-III gene *(HCN1)* and four stage-IV genes *(NELL1, ZNF135, FAM123A, LAMA1)*. Each of these stage-salient genes presented an |lfc M-value| > 0.4 with respect to the other stages, validating their salience. Figures 15,16 represent boxplots of the consensus biomarkers, and Table 5 presents a summary of the consensus analysis. Gene ontology (GO) analysis [40] of the consensus biomarkers yielded processes related to structural integrity of cell division processes, immunity dysfunction, and cell migration (Table 6). Detailed GO results are presented in the Supplementary Information S12.

**Table 5.** Stage-salient biomarkers. The results of the consensus analysis and univariate survival analysis are summarized.

| HGNC ID | Gene Name | Methods providing identical results | Stage salience | Status of methylation | Inferred effect on gene expression | P-values from modelling |  | P-values from univariate survival analysis |  |
| --- | --- | --- | --- | --- | --- | --- | --- | --- | --- |
|  |  |  |  |  |  | M value | Averep | Cox analysis | Kaplan Meier |
| 3603 | FBN1 | Averep, CHAMP | I | Hyper | Down | 0.310 | 0.040 | 0.036 | 0.025 |
| 3811 | FOXG1 | Averep, Mvalue, CHAMP, Methylmix | II | Hyper | Down | 3.25E-16 | 0.003 | 0.019 | 0.037 |
| 4845 | HCN1 | Averep, Mvalue, CHAMP | III | Hyper | Down | 1.32E-17 | 0.022 | 0.031 | 0.059 |
| 7756 | NELL1 | Mvalue, CHAMP, Methylmix | IV | Hyper | Down | 2.49E-68 | 0.0614 | 0.283 | 0.27 |
| 12919 | ZNF135 | Mvalue, CHAMP, Methylmix | IV | Hyper | Down | 1.29E-76 | 0.0622 | 0.096 | 0.084 |
| 26360 | FAM123A | Mvalue, CHAMP, Methylmix | IV | Hyper | Down | 5.09E-115 | 0.0966 | 0.30 | 0.28 |
| 6481 | LAMA1 | Mvalue, CHAMP, Methylmix | IV | Hyper | Down | 7.57E-86 | 0.297 | 0.052 | 0.051 |

**Table 6.** GO analysis of stage-salient genes in the order of decreasing significance (i.e, increasing p – value). Ontology: Cellular Compartment (CC), Molecular Function (MF), Biological Process (BP).

| GO ID | Term | Ontology |  |  | ep-value |
| --- | --- | --- | --- | --- | --- |
| GO:1990047 | spindle matrix | CC |  |  | 0.000148 |
| GO:0030109 | HLA-B specific inhibitory MHC class I receptor activity | MF |  |  | 0.000297 |
| GO:0032396 | inhibitory MHC class I receptor activity | MF |  |  | 0.000594 |
| GO:0042609 | CD4 receptor binding | MF |  |  | 0.001187 |
| GO:0032393 | MHC class I receptor activity | MF |  |  | 0.001336 |
| GO:0050930 | induction of positive chemotaxis | BP |  |  | 0.001632 |
| GO:0050927 | positive regulation of positive chemotaxis | BP |  |  | 0.003263 |
| GO:0050926 | regulation of positive chemotaxis | BP |  |  | 0.003411 |
| GO:0008608 | attachment of spindle microtubules to kinetochore | BP |  |  | 0.004299 |
| GO:0007094 | mitotic spindle assembly checkpoint | BP |  |  | 0.004448 |

**Figure 15.**
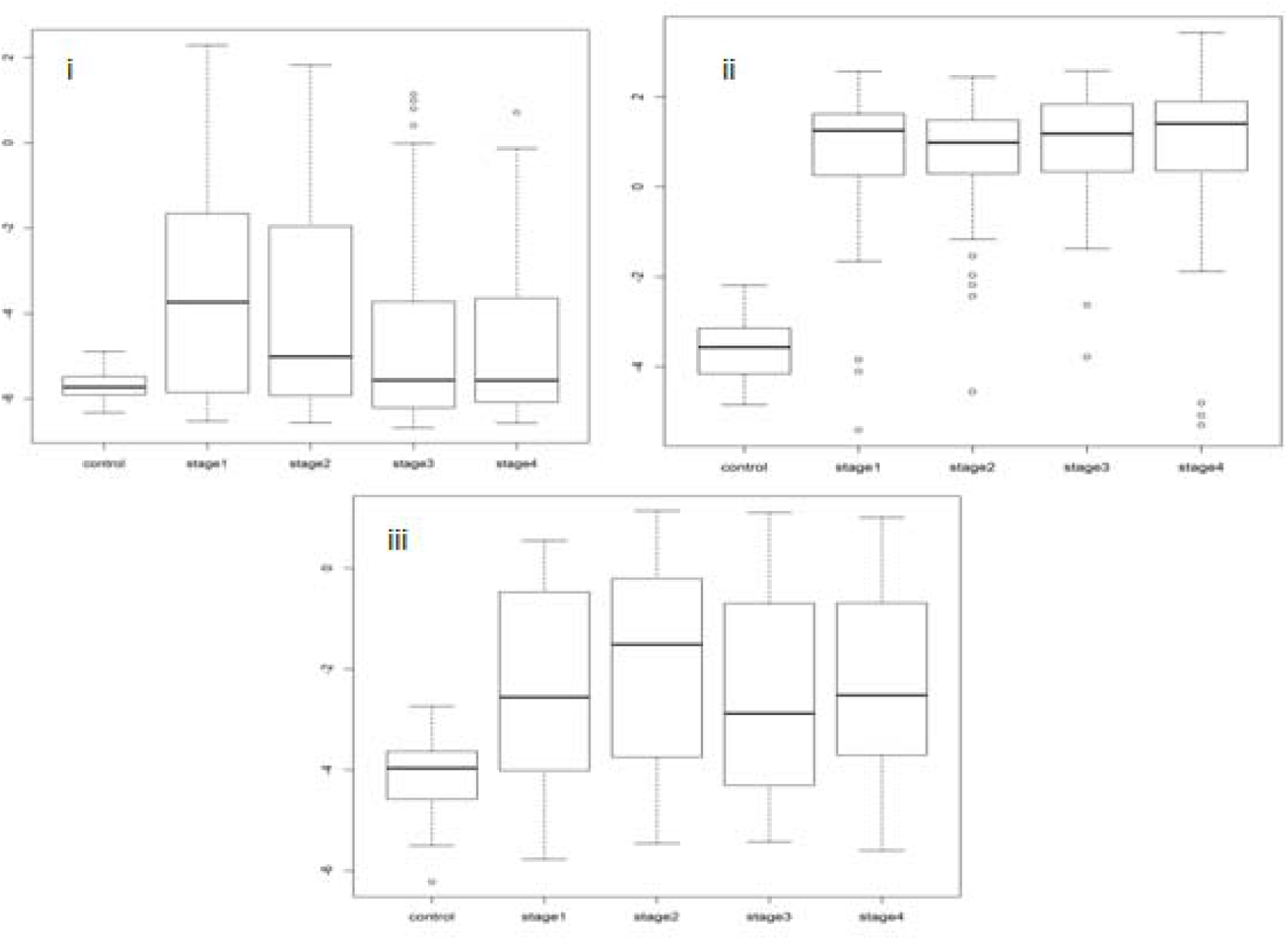
Boxplot analysis of stage-salient genes. (i) Stage-I Gene FBN1, (ii) Stage-II Gene – FOXG1, (iii) Stage-III Gene – HCN1.

**Figure 16.**
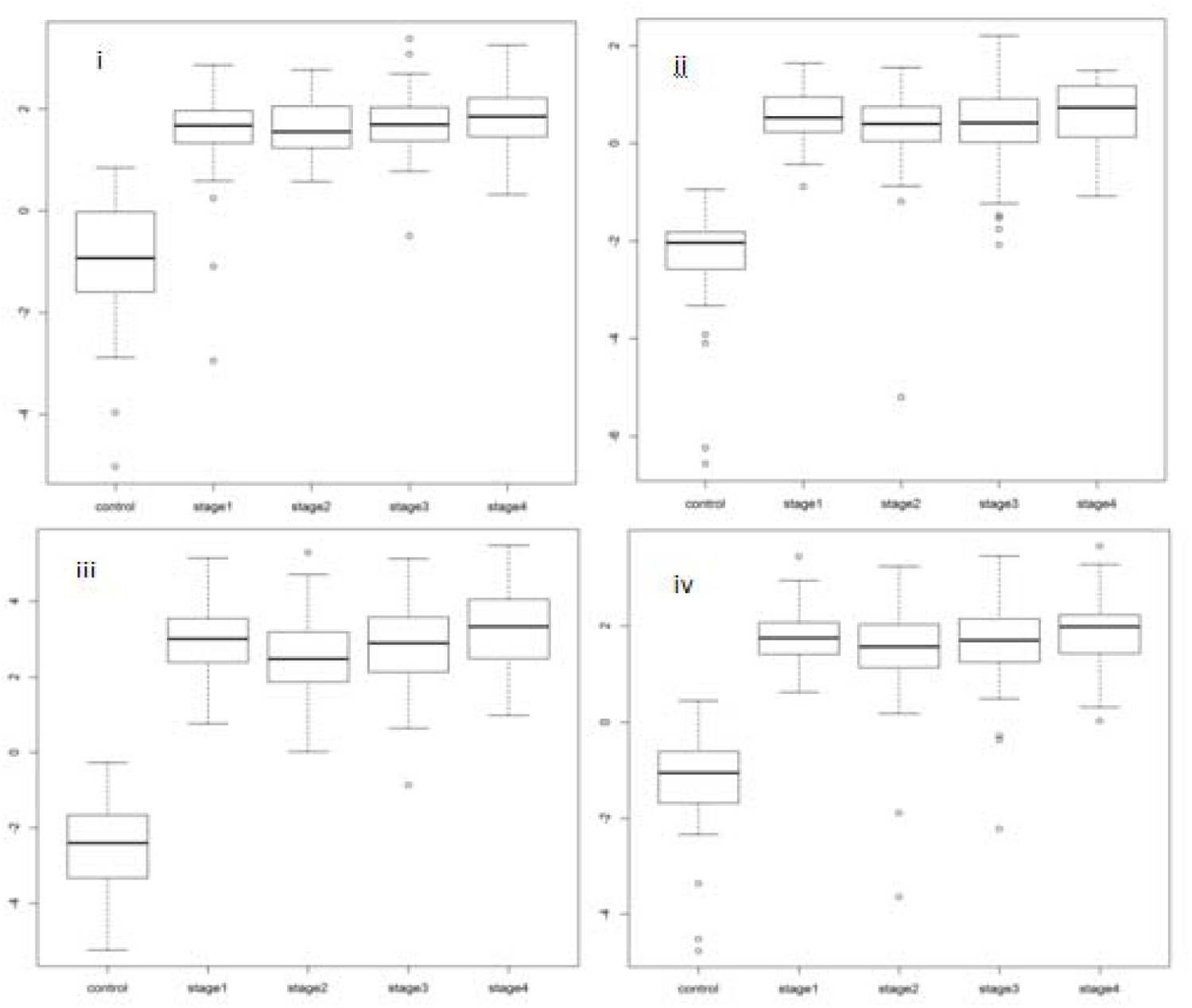
Boxplot representation of stage-wise methylation levels for Stage-IV salient genes. (i) LAMA1, (ii) NELL1, (iii) FAM123A, (iv) ZNF135.

### Survival analysis

We constructed independent prognostic models of the stage-salient genes and the corresponding univariate Kaplan-Meier plots of prognostically significant biomarkers are shown in Fig. 17. These include FBN1, FOXG1, HCN1, and LAMA1. Rational combinations of stage-salient genes, termed ColoRectal cancer Signatures (CRS), were modelled using multivariate Kaplan-Meier regression, to yield a risk score. Risk scores were then used to estimate survival-effect significance, as described in Methods. We found that CRS12 signature (consisting of stages I and II biomarkers: FBN1, FOXG1) yielded significant risk scores in the multivariate Kaplan-Meier analysis, and both CRS12 and CRS34 (which consisted of stages III and IV biomarkers: HCN1, NELL1, ZNF135, FAM123A, LAMA1) were significant in estimating overall survival (prognosis p-value ≤ 0.02) (Figure 18). The results of the survival analysis are summarised in Table 7. Supplementary Information S13 provides survival plots of all possible signatures; it is observed that the optimal signatures immediately yield an early-stage panel (CRS12), and a late-stage panel (CRS34)..

**Table 7.**
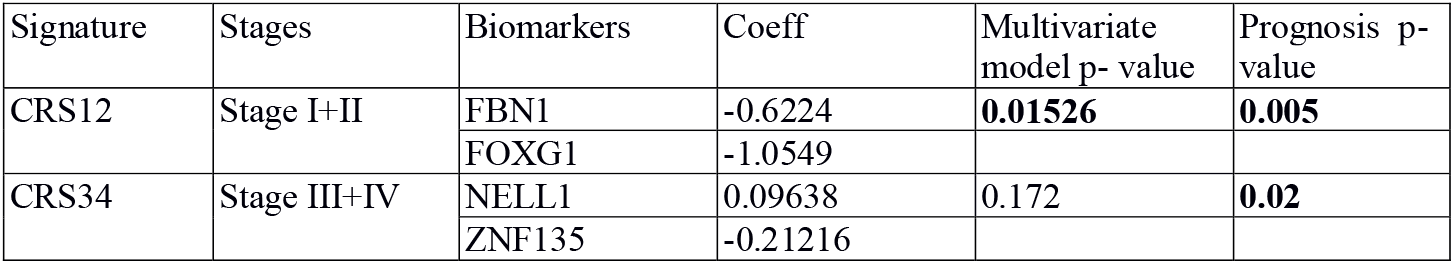
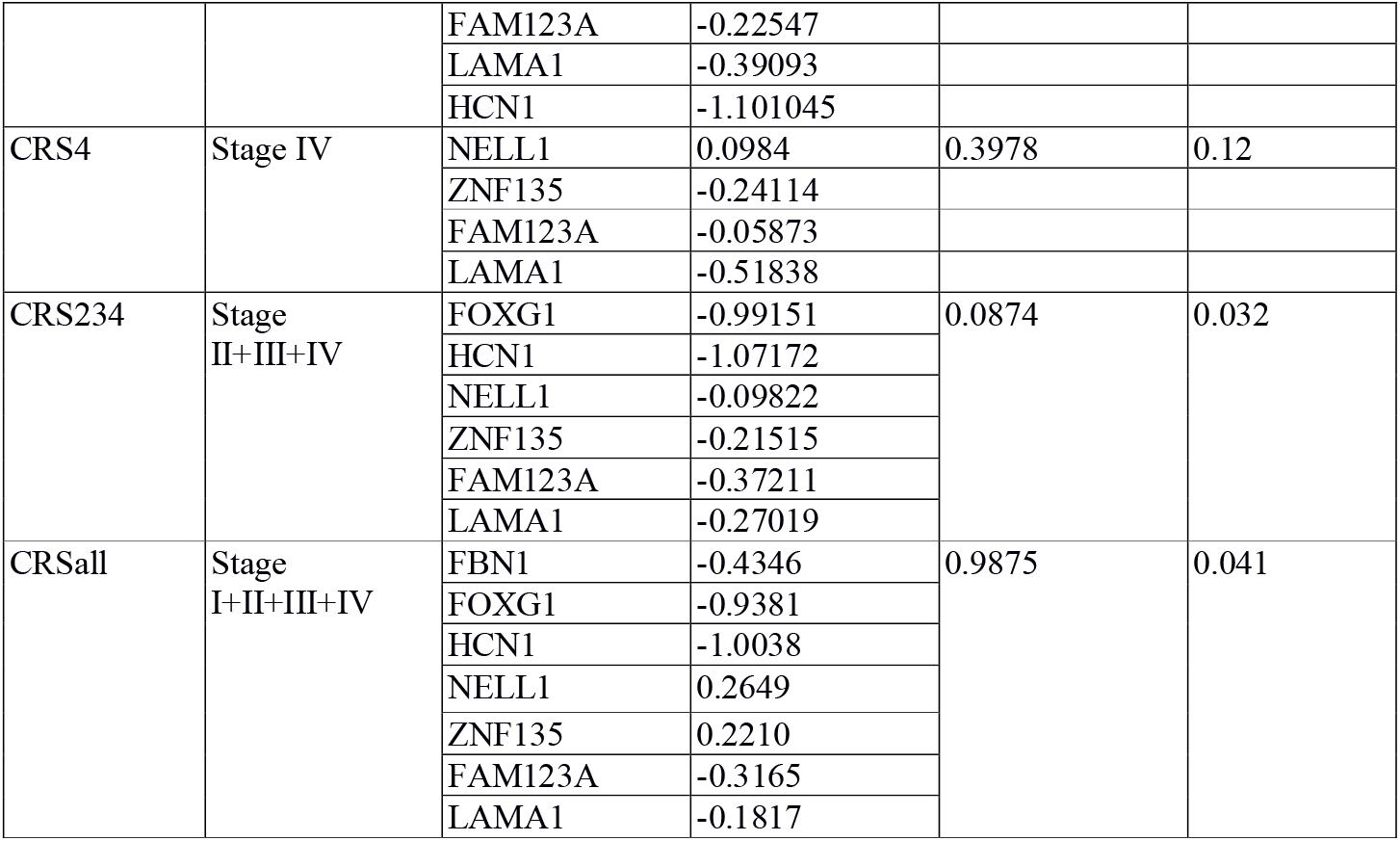
Summary of the multivariate prognostic models. Significant signatures are emphasized.

| Signature | Stages | Biomarkers | Coeff | Multivariate model p- value | Prognosis p- value |
| --- | --- | --- | --- | --- | --- |
| CRS12 | Stage I+II | FBN1 | -0.6224 | **0.01526** | **0.005** |
|  |  | FOXG1 | -1.0549 |  |  |
| CRS34 | Stage III+IV | NELL1 | 0.09638 | 0.172 | **0.02** |
|  |  | ZNF135 | -0.21216 |  |  |
|  |  | FAM123A | -0.22547 |  |  |
|  |  | LAMA1 | -0.39093 |  |  |
|  |  | HCN1 | -1.101045 |  |  |
| CRS4 | Stage IV | NELL1 | 0.0984 | 0.3978 | 0.12 |
|  |  | ZNF135 | -0.24114 |  |  |
|  |  | FAM123A | -0.05873 |  |  |
|  |  | LAMA1 | -0.51838 |  |  |
| CRS234 | Stage II+III+IV | FOXG1 | -0.99151 | 0.0874 | 0.032 |
|  |  | HCN1 | -1.07172 |  |  |
|  |  | NELL1 | -0.09822 |  |  |
|  |  | ZNF135 | -0.21515 |  |  |
|  |  | FAM123A | -0.37211 |  |  |
|  |  | LAMA1 | -0.27019 |  |  |
| CRSall | Stage I+II+III+IV | FBN1 | -0.4346 | 0.9875 | 0.041 |
|  |  | FOXG1 | -0.9381 |  |  |
|  |  | HCN1 | -1.0038 |  |  |
|  |  | NELL1 | 0.2649 |  |  |
|  |  | ZNF135 | 0.2210 |  |  |
|  |  | FAM123A | -0.3165 |  |  |
|  |  | LAMA1 | -0.1817 |  |  |

**Figure 17.**
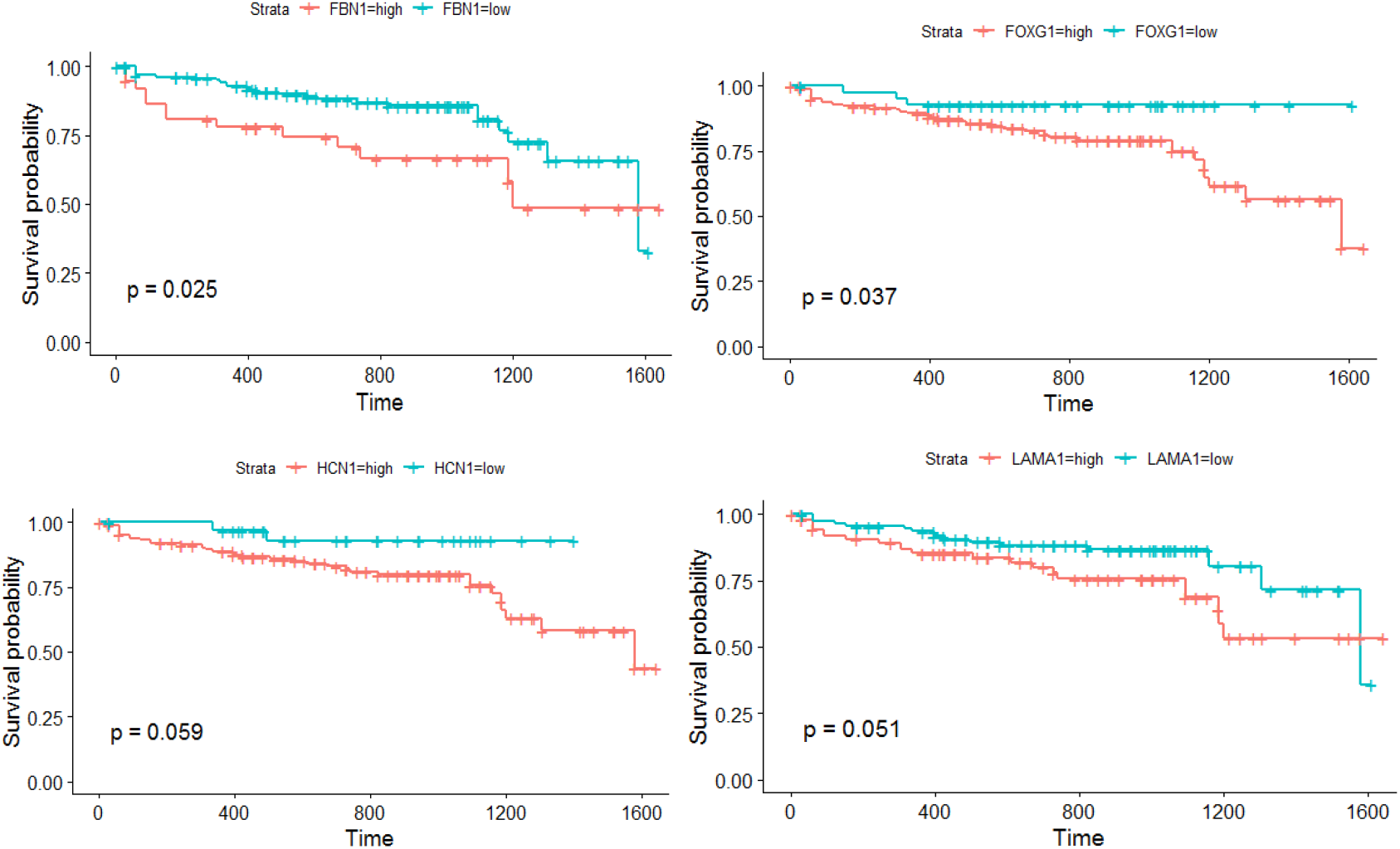
K-M plots for the prognostically significant stage-salient genes. (A) FBN1, (B) FOXG1, (C) HCN1, and (D) LAMA1.

**Figure 18.**
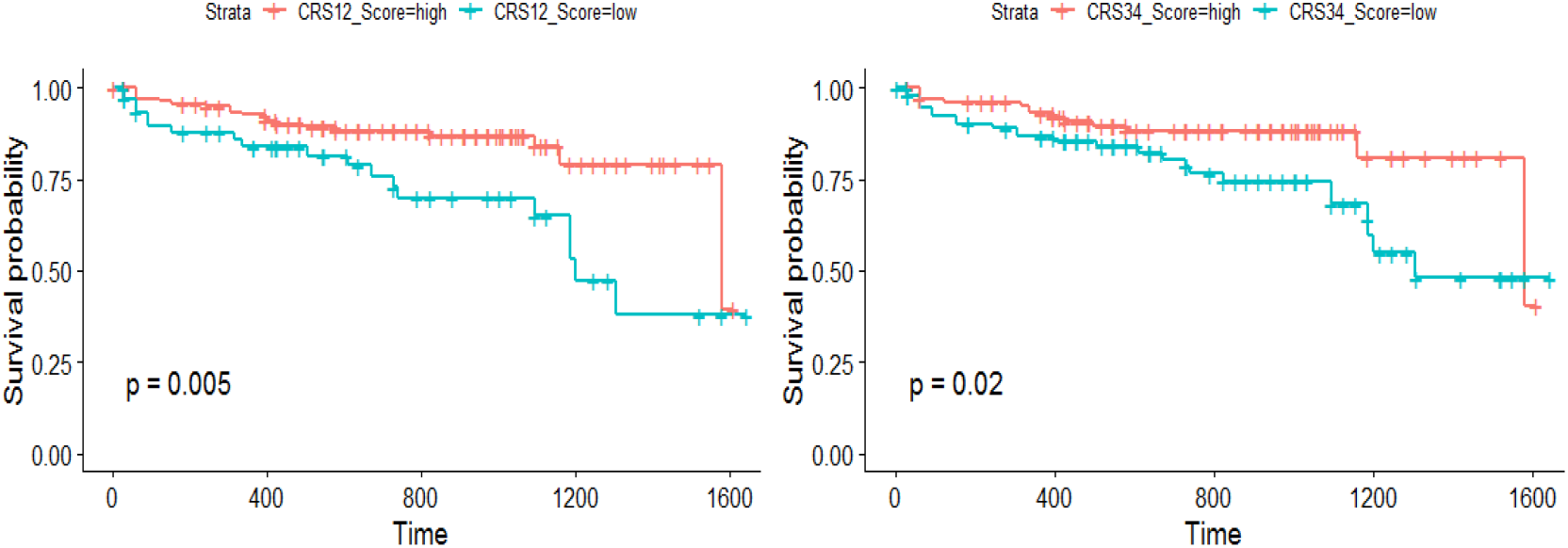
Survival analysis of combination biomarker panels shows significance. (A) Early-stage panel; and (B) Late-stage panel.

## DISCUSSION

CRC development is due to the accumulation of genetic and epigenetic changes of which DNA methylation is of prime importance. DNA methylation profiles of colorectal cancer have been investigated in several previous studies using various approaches [41, 42]. It is well-known that changes in methylation status correspond with CRC progression [43]. Here we have designed a comprehensive approach to systematically analyze stage-differentiated DNA methylation patterns in colorectal cancer and their relationship to patient survival. Our study has yielded consensus stage-salient significantly differentially methylated genes, stage-agnostic genes, and their prognostic value. A total of seven genes were identified by at least two methods, and of those, six were identified by at least three methods (FBN1 being the exception). None of the stage-salient genes is included as a cancer gene or hallmark gene in the Cancer Gene Census [44], while HCN1 alone is reported as a candidate cancer gene based on mouse insertional mutagenesis experiments [45]. Below, a discussion of all the stage-salient DMGs (Table 5) is provided with respect to the existing literature.

### Early-stage salient DMGs

Promoter hypermethylation of FBN1, a glycoprotein component of calcium-binding extracellular matrix microfibrils [46], is a recognized biomarker of CRC [47, 48]. Our analysis supports this literature, while pinpointing the stage I-salience in its action. FOXG1 is well-known as an etiological factor in certain neurological disorders and plays a role in the epithelial-mesenchymal transition of CRC cells (a key hallmark of cancer progression), and is known to be overexpressed in CRC patients [49]. It is a nodal gene, with connections to oncogenic pathways like WNT pathway in hepatocellular carcinoma [50] and TGF-β pathway in ovarian cancer [51]. Interestingly, FOXG1 was found to be a hypermethylated stage-II salient gene. HCN1, coding for hyperpolarization-activated cyclic nucleotide-gated channel subunits is associated with low survival rates in breast, brain, and colorectal cancer [52]. We have identified HCN1 as a stage-III hypermethylated gene, suggesting a loss-of-function mechanism for its tumorigenic potential.

### Stage-IV salient DMGs

Our study has provided clear evidence that hypermethylation of LAMA1 (which codes for α-laminin of the extracellular matrix) is a stage IV-specific signature. Experimental evidence for the hypermethylation of the promoter region of LAMA1 in CRC patients is available [53]. NELL1 is a known tumor suppressor gene [54], whose hypermethylation is associated with poor survival outcomes [55]. Here it is found to be a stage IV-specific hypermethylated gene, resonating with the above findings. ZNF135 is involved in regulation of cell morphology and cytoskeletal organizations, and its expression and epigenetic regulation have been reported to be key in cancers of the cervix and esophagus, respectively [56, 57]. Here we have found that epigenetic silencing of ZNF135 is a key feature of stage-IV CRC. FAM123A, also known as AMER2, is associated with microtubule proteins [58], and is a lesser known cousin of FAM123B, a tumor-suppressor whose loss-of-function by mutation, methylation and copy-number aberrations is known play pivotal roles in colorectal cancer, especially in older patients [59,60,61]. It is significant that our study has uncovered FAM123A as a hypermethylated stage IV-specific DMG, signalling the need for experimental investigations. There is very little literature on the cancer significance of any of the above stage-salient genes, marking our findings as novel and important in the context of gaps in our knowledge.

### Putative CIMP signature

Aberrant methylation of CpG promoter regions causes stable repression of transcription leading to gene-silencing [62,63]. In the context of tumorigenic processes, this is likely to lead to loss-of-function of tumor-suppressor genes. Multiple CpG islands might be methylated simultaneously in some cancers, paving the way for CpG island methylator phenotype (CIMP), first discovered in colorectal cancer [64]. CIMP is characterised by hypermethylation of CpG islands surrounding the promoter regions of genes involved in cancer onset and progression [65]. The phenotype is heterogenous with the type of tumor [66] and dependent on definition [67]. In this background, it is less straightforward to interpret the functional importance of hypermethylation of individual genes. Still it is clear from Table 5 that the stage-salient hypermethylated biomarkers identified in our study could constitute an aggregate novel CIMP. The original CIMP had been associated with advanced T staging (T3/T4) [68], which accords with our finding of five hypermethylated stage IV-salient DMGs. Epigenetic intervention for CIMP-positive cancers has been suggested as a possible treatment strategy [69].

The biomarkers contributing to the putative CIMP were tested with Cox regression and then evaluated independently as well as in combination for prognostic significance. Five of the seven stage-salient genes were prognostically significant in both the Cox univariate model and the Kaplan-Meier analysis (Table 5). A multivariate analysis of biomarker panels uncovered two signatures, an early-stage CRS12, and a late-stage CRS34 that might be prognostically valuable. In particular, CRS12 suggests a significant early-stage biomarker panel (p-value < 0.01) for the effective prognosis and stage-sensitive detection of colorectal cancer.

The current standard of CRC screening is colonoscopy, an invasive method with a significant rate of complications. A non-invasive method based on molecular diagnostics would improve patient satisfaction and efficiency. Several studies have been conducted to identify and/or validate biomarkers for CRC diagnosis. It is recognized that DNA methylation patterns could serve as valid biomarker candidates [70,71]. Freitas et al., have validated the performance of a 3-gene biomarker panel for the detection of colorectal cancer irrespective of the molecular subtype [72]. However optimal stage-salient epigenetic biomarkers have not yet been reported. Using hypermethylated DNA patterns as cancer markers offers the advantage of providing small targets with high concentrations of CpG for assays, useful for the design of analytical amplicons [73]. Hypermethylation in gene body and upstream control regions like enhancers and insulators might affect transcription differently than hypermethylation of promoter regions [74.75]. Further DNA methylation patterns in noncoding RNA genes seem to be important in tumorigenesis and progression [76]. Non-encoding RNAs themselves play a significant role in epigenetic modification through the phenomenon of RNA-directed DNA methylation [77]. The nuanced relationship between methylation and gene transcription does urge the interpretation of our results with caution, contingent on experimental validation, however consensus study designs such as ours suffer less uncertainties with respect to the results. Since methylation is a direct, ubiquitous and effective mechanism of epigenetic regulation used by plants and animals [78], it is hoped that our studies would advance our understanding of the complex effects of methylation events, patterns, and landscapes in different scenarios, including in the developmental stages of life.

## CONCLUSION

We have developed a comprehensive computational framework for the consensus identification of stage-differentiated significant differentially methylated genes, and evaluation of their prognostic significance. Our analysis has yielded seven stage-salient genes, all hitherto unreported in the literature: one stage-I gene *(FBN1)*, one stage-II gene *(FOXG1)*, one stage-III gene *(HCN1)* and four stage-IV genes *(NELL1, ZNF135, FAM123A, LAMA1)*. Stage-salient genes could serve as diagnostic biomarkers. The top stage-agnostic genes could serve as targets for drug discovery in CRC therapy. All the stage-salient genes were found to be hypermethylated, indicating a novel CIMP-like character possibly promoting epigenetic destabilisation that merits further investigation. Independent prognostic evaluation of the stage-salient genes yielded significance for FBN1, FOXG1, HCN1, and LAMA1. Survival analysis of biomarker signatures composed of the stage-salient genes yielded a significant early-stage panel and a significant late-stage panel. Robust consensus approaches, like the one used here, are more reliable, and the epigenetic biomarkers identified in our study could greatly advance the early detection of colorectal cancers, their treatment and prognostic evaluation. Our approach is extendable to the investigation of epigenomics in other cancers, normal/disease conditions, and perhaps even developmental biology.

## Data Availability

All data and results are available as available as supplementary information:
https://doi.org/10.6084/m9.figshare.13013852

## ACKNOWLEDGMENTS

We are grateful to the School of Chemical and BioTechnology, SASTRA Deemed University for computing and infrastructure support. A.P. would like to acknowledge funding from DST-SERB grant EMR/2017/000470/BBM.

## AUTHOR CONTRIBUTIONS

A.P. conceived, designed and supervised the work. S.M. and A.R. performed the research; A.P., S.M., and A.R. analyzed and interpreted the results. A.P. wrote the manuscript. All authors approved the manuscript.

